# Integrative approach identifies *SLC6A20* and *CXCR6* as putative causal genes for the COVID-19 GWAS signal in the 3p21.31 locus

**DOI:** 10.1101/2021.04.09.21255184

**Authors:** Silva Kasela, Zharko Daniloski, Tristan X. Jordan, Benjamin R. tenOever, Neville E. Sanjana, Tuuli Lappalainen

## Abstract

To date the locus with the most robust human genetic association to COVID-19 susceptibility is 3p21.31. Here, we integrate genome-scale CRISPR loss-of-function screens and eQTLs in diverse cell types and tissues to pinpoint genes underlying COVID-19 risk. Our findings identify *SLC6A20* and *CXCR6* as putative causal genes that mediate COVID-19 risk and highlight the usefulness of this integrative approach to bridge the divide between *correlational* and *causal* studies of human biology.

## Background

COVID-19, which is caused by SARS-CoV-2 (Severe Acute Respiratory Syndrome-Coronavirus-2) infection, results in diverse individual disease courses from asymptotic carriers to severe disease with respiratory failure [1, 2]. A growing body of evidence suggests also a notable role of genetic factors in COVID-19 susceptibility and severity [3–5], and several studies have identified a robust association at chromosome 3p21.31 locus. However, the functional mechanisms of this association are unclear. The locus includes multiple protein-coding genes, for example, *LIMD1, SACM1L, SLC6A20, LZTFL1, CCR9, FYCO1, CXCR6*, and *XCR1*, many of which have potentially relevant role in the pathophysiology of COVID-19. As genetic variants have been shown to often exert their effect on complex traits or disease via *cis*-regulation of gene expression [6, 7], expression quantitative trait locus (eQTL) mapping could serve as a mean to pinpoint candidate genes for trait or diseases of interest.

We previously performed a genome-scale CRISPR loss-of-function screen to identify genes required for SARS-CoV-2 viral infection in human lung epithelial-like cells expressing *ACE2* (A549^ACE2^) [8]. In this work, we present an integrative approach using the results of the CRISPR screen and eQTLs in various cell types and tissues from the eQTL Catalogue [9] and Genotype Tissue Expression (GTEx) [7] v8 data release to pinpoint genes underlying COVID-19 risk in the chr3 locus. We show that genes enriched in the *in vitro* CRISPR screen contribute to COVID-19 risk in humans, and highlight *SLC6A20* and *CXCR6* as the putative causal genes in the chr3 COVID-19 risk locus.

## Results and Discussion

We sought to determine to what extent the top-ranked genes from the CRISPR screen — whose loss reduces SARS-CoV-2 infection — mediate disease risk in humans by leveraging data from COVID-19 genome-wide association study (GWAS). We hypothesized that genetic regulatory variants for genes pinpointed by the CRISPR screen would show an increased signal for genetic association for COVID-19 in the human population. To this end, we analyzed if eQTLs from lung tissue [7] for these genes would be enriched for overall association signal in data from the COVID-19 Host Genetic Initiative [4] with six COVID-19-related GWAS (release 3) with different definition of cases and controls. Indeed, we observed inflation of the GWAS association signal for variants that are *cis*-eQTLs for top-ranked CRISPR hit genes in the hospitalized vs not hospitalized COVID-19 GWAS, compared to all lung eQTLs (λ_0.1_ = 2.14, permutation *P*-value = 0.046, **Fig. 1a-b**). We did not see a significant signal for the other five GWAS, which could be explained by the low power of these early GWAS data (**Additional file 1: Figure S1**). Nevertheless, our results establish the link between genes enriched in the *in vitro* CRISPR screen and the genetic component of COVID-19 risk in humans.

**Fig. 1.**
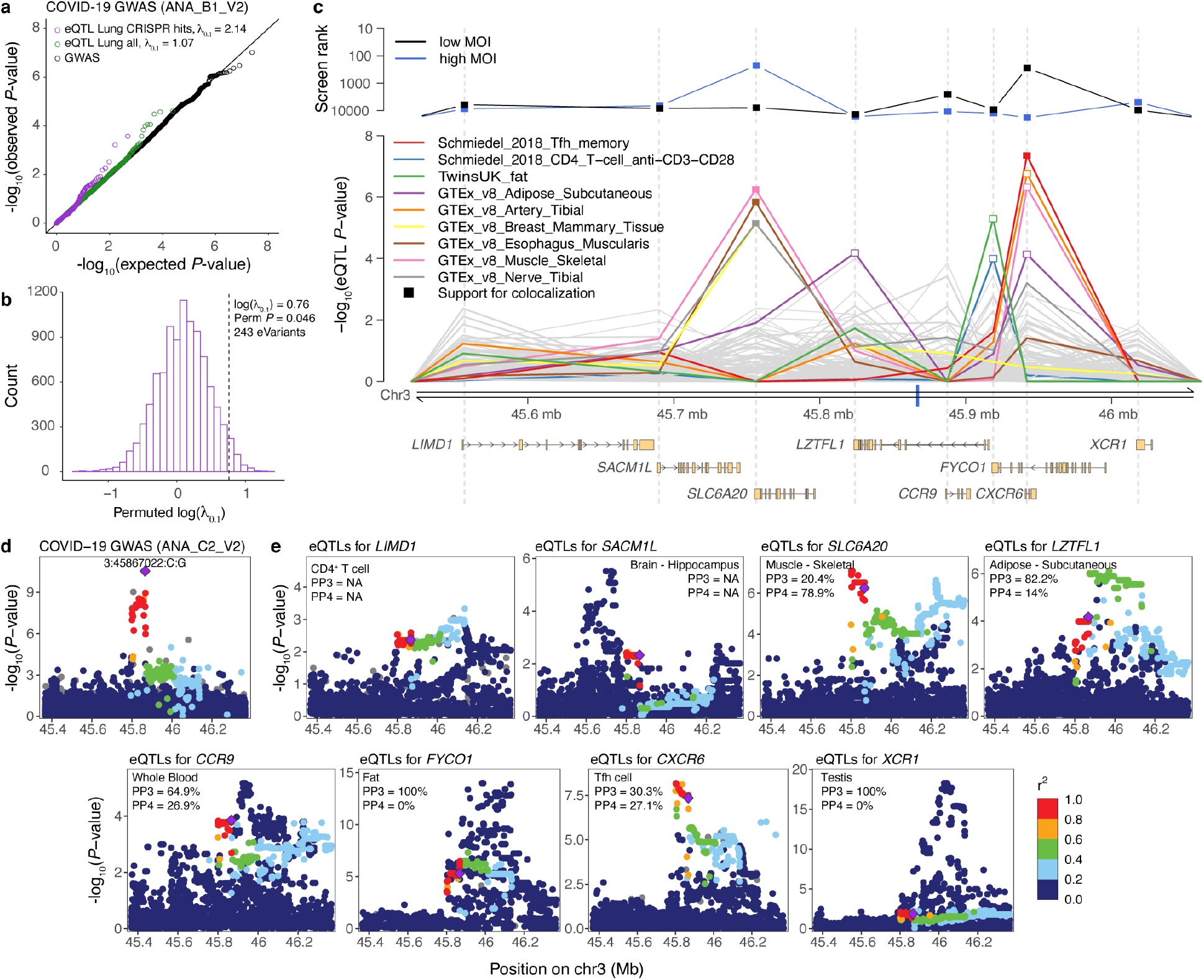
Genetic regulatory effects of CRISPR hit genes and prioritization of genes in the 3p21.31 cluster associated with COVID-19 GWAS. **a**, Q-Q plot showing the expected and observed *P*-value distribution of hospitalized vs not hospitalized COVID-19 GWAS (ANA_B1_V2) for three sets of variants: all variants tested in GWAS (black), variants that are lead eQTLs in GTEx Lung (green), and variants that are lead eQTLs in GTEx Lung for the top-ranked genes enriched in at least one CRISPR screen (purple). Inflation estimate λ_0.1_ measures the inflation of test statistics relative to the chi-square quantile function of 0.1. **b**, Histogram of the permuted log(λ_0.1_) to test the significance of the inflation of variants that are eQTLs for CRISPR screen hit genes (*n* = 243) in Lung in ANA_B1_V2 COVID-19 GWAS. **c**, Prioritization of genes in the 3p21.31 locus associated with COVID-19 vs population (ANA_C2_V2) GWAS. The top panel shows the ranking of the genes in the locus according to the second-best guide RNA score in the low MOI (black) and high MOI (blue) pooled CRISPR screens. The middle panel shows the eQTL *P*-values for the lead GWAS variant 3:45867022:C:G (denoted as a blue tick on the *x*-axis) in different cell types and tissues from the eQTL Catalogue and GTEx v8, 110 eQTL data sets in total. Highlighted are nine cell types/tissues, where the eQTL *P*-value for the lead GWAS variant is < 10^−4^ for at least one gene in the region. Filled square denotes suggestive support for colocalization between the GWAS and eQTL signal (posterior probability for one shared causal variant (PP4) > 0.25). The bottom panel depicts the transcripts of the eight protein-coding genes in the locus. Ranks and eQTL *P*-values are aligned to match the start of the gene which is shown as a grey dashed line across the panels. **d-e**, Regional association plots of the COVID-19 vs population (ANA-C2-V2) GWAS (**d**) and *cis*-eQTLs for the eight genes (**e**) from the associated locus in the cell type/tissue where the lead GWAS variant has the lowest eQTL *P*-value. Purple diamond denotes the lead GWAS variant, and the data points are colored based on the (weighted average) LD between the lead GWAS variant and other variants in the region in the respective study population. PP3 and PP4 - posterior probability for two different variants or one shared causal variant in coloc, respectively.

To elucidate which genes in the chromosome 3 locus might mediate the genetic association, we first observed that Solute Carrier Family 6 Member 20 (*SLC6A20*) gene and C-X-C Motif Chemokine Receptor 6 (*CXCR6*) gene have a relatively high rank in our CRISPR screen (**Fig. 1c**). Next, we analyzed if the COVID-19 ANA_C2_V2 GWAS lead variant 3:45867022:C:G affects expression of any of these genes. Using data from the eQTL Catalogue [9] and GTEx v8 [7], we analyzed all eQTL associations for this variant, and performed colocalization analysis [10, 11] to assess if the eQTL and GWAS signal share a genetic cause. *CXCR6* and *SLC6A20* stood out, with the eQTL data indicating that the COVID-19-associating variant affects the expression of these two genes (**Fig. 1c, Additional file 1: Figure S2, Additional file 2: Table S1**). The eQTL affecting *CXCR6* is active in memory T follicular helper (Tfh) CD4^+^ T cells [12] and has a suggestive colocalization with the GWAS (**Fig. 1d-e**). The colocalization signal was very strong for the *cis*-eQTL for *SLC6A20* in four tissues from GTEx - esophagus muscularis, breast mammary tissue, skeletal muscle, and tibial nerve (**Additional file 1: Figure S3**). Other genes in the region, such as *LZTFL1, FYCO1*, and *XCR1*, also have regulatory variants affecting their expression, but two distinct variants likely drive the GWAS and eQTL signals (**Fig 1d-e**). Given that both the CRISPR screen and eQTL data support the causal role of *SLC6A20* and *CXCR6*, it is possible that the COVID-19 GWAS association in the 3p21.31 locus is driven by pleiotropic effects of the same variant on multiple genes in different cell types.

In addition to human genetic evidence and support from experimental data, both *SLC6A20* and *CXCR6* have a plausible biological function that could affect COVID-19. Notably, SLC6A20 functionally interacts with ACE2 [13], the receptor of the SARS-CoV-2 Spike protein that is the key host gene for viral entry [14]. In contrast, CXCR6 regulates the localization of resident memory T (T_RM_) cells in the lung and maintains a pool of airway T_RM_ cells, critical for cellular immunity against respiratory pathogens [15]. It is also an alternate coreceptor for HIV [16, 17], raising a hypothesis of a similar function in lung cells for SARS-CoV-2. The fact that we observed eQTLs for *SLC6A20* and *CXCR6* in different tissues and cell types other than lung highlights the complexity and possibility of extrapulmonary spread of SARS-CoV-2 [18], as well as potential pleiotropic effects of these genes in multiple cell types and physiological processes leading to COVID-19. Importantly, GWAS can only show associations for common variants in the human population, and thus, the functional role of these genes in lung cells, captured by the screen, may not be captured by GWAS and eQTL data. Nevertheless, corroborating evidence from entirely independent approaches pinpointing *SLC6A20* and *CXCR6* warrants further studies exploring the virological or mechanistic mode of action of these two genes.

## Conclusion

In this work, we demonstrated that genes required for SARS-CoV-2 infection *in vitro* also contribute to COVID-19 susceptibility in humans. Our integrative analysis of the 3p21.31 GWAS locus indicated *SLC6A20* and *CXCR6* as potential protein-coding genes through which noncoding variants associated with COVID-19 risk in human patients may function. This integrative approach should prove useful for other human diseases and pathogens to bridge the divide between *correlational* and *causal* studies of human biology.

## Methods

### Genome-wide CRISPR screen and guide RNA analysis

Details regarding the SARS-2 CRISPR screen in A549^ACE2^ human lung epithelial cells that over-express ACE2 (A549^ACE2^) have been described before [8]. Briefly, human GeCKOv2 A and B libraries (Addgene, 1000000048) were used for the genome-wide CRISPR-Cas9 screen [19]. After transduction and selection for the GeCKOv2 library (maintaining ∼1000-fold library representation), we infected 125 million A549^ACE2^ cells with SARS-CoV-2 virus (NIAID BEI isolate USA-WA1/2020) at two multiplicity of infections (MOIs): 0.01 (low MOI) and 0.3 (high MOI), which differ by the amount of virus used to infect the cells. Surviving cells were collected on day 6 post-infection, when genomic DNA was isolated, guide RNAs were recovered by PCR, and guide representation was determined by next generation sequencing (Illumina).

We aligned trimmed sequencing reads to the GeCKOv2 reference using bowtie v1.1.2 [`-a -- best --strata -v 1 –norc`] allowing 1 nucleotide mismatch to determine the read counts per guide. Alignment rates were ∼80% for all samples. We normalized read counts between biological samples and computed a fold-change by comparison of SARS-CoV-2-infected samples to the uninfected control. Genes were then ranked based on the second-best guide as described before [20–22]. Briefly, each gene in the human GeCKOv2 A+B library is targeted by 6 different guides. Genes are ranked based on the guide RNA with second highest fold-change (sbScore).

### Inflation of COVID-19 GWAS signal for variants that are *cis*-eQTLs

To estimate the importance of CRISPR screen hit genes in susceptibility to COVID-19 in humans, we tested if we observe inflation of COVID-19 GWAS signal for variants that are eQTLs in GTEx v8 lung [7] for top-ranked genes enriched in at least one CRISPR screen (*n* = 588 genes). We used summary statistics of the six COVID-19 GWAS generated by the COVID-19 Human Genetics Initiative (HGI) [4] (round 3 meta-analyses, released July 2, 2020): 1) ANA_A2_V2 - very severe respiratory confirmed covid vs. population, 2) ANA_B1_V2 - hospitalized covid vs. not hospitalized covid, 3) ANA_B2_V2 - hospitalized covid vs. population, 4) ANA_C1_V2 - covid vs. lab/self-reported negative, 5) ANA_C2_V2 - covid vs. population, 6) ANA_D1_V2 - predicted covid from self-reported symptoms vs. predicted or self-reported non-covid. We measured inflation using the lambda inflation statistic relative to the chi-square quantile function of 0.1, λ_0.1_, *i*.*e*., estimating the inflation among 10% of the most significant tests. We calculated λ_0.1_ for two sets of eQTLs: 1) all lead eQTLs in lung (14,113 genes with significant eQTLs in lung at FDR 5%, serving as a background set), 2) all lead eQTLs in lung for CRISPR hit genes (274 out 588 top-ranked genes have eQTLs in lung at FDR < 0.05).

To test the significance of λ_0.1_, we used a permutation-based test. We selected *n* number of lead eQTLs from the background set *k* = 10,000 times, where *n* is the number of lead eQTLs for CRISPR hit genes tested in a given COVID-19 GWAS, and calculated λ_0.1_ on the permuted data. To calculate two-sided permutation *P*-values, we applied log-transformation on the permuted λ_0.1_ values to get a symmetric null distribution. We then calculated permutation *P*-value as the proportion of permuted log(λ_0.1_) as extreme as or more extreme than the observed log(λ_0.1_).

### Prioritization of genes in the 3p21.31 locus based on CRISPR screen and eQTL data

We focused on eight genes, *LIMD1, SACM1L, SLC6A20, LZTFL1, CCR9, FYCO1, CXCR6*, and *XCR1*, in the 3p21.31 locus that has been shown to robustly associate with COVID-19 susceptibility [3–5]. Firstly, we ranked the genes based on the sbScore in the CRISPR screen. Secondly, we gathered summary statistics for 110 eQTL data sets from the eQTL Catalogue [9], (61 data sets, mostly immune cell types with and without treatment) and GTEx v8 [7] (49 tissues). We prioritized the genes based on the *P*-value in eQTL studies for the lead GWAS variant 3:45867022:C:G from the covid vs population (ANA_C2_V2, largest COVID-19 GWAS to date with 6,696 total cases and 1,073,072 total controls) GWAS generated by the COVID-19 HGI. Since observing low eQTL *P*-value for the lead GWAS variant does not necessarily translate into shared genetic causality, we further performed colocalization analysis.

### Colocalization analysis of COVID-19 GWAS and *cis*-eQTLs in the 3p21.31 locus

To assess evidence for shared causal variant of a *cis*-eQTL and a COVID-19 GWAS, we used the Bayesian statistical test for colocalization, coloc [10], assuming one causal variant per trait. We only included *cis*-eQTLs for genes for colocalization test, if there was a *cis*-eQTL with a nominal *P*-value < 10^−4^ within 100kb of the lead ANA_C2_V2 COVID-19 GWAS variant. Coloc was run on a 1Mb region centered on the lead GWAS variant (+/-500 kb from the variant) with prior probabilities set to *p*_*1*_ = 10^−4^, *p*_*2*_ = 10^−4^, *p*_*3*_ = 5×10^−6^. We defined suggestive support for colocalization between the ANA_C2_V2 COVID-19 GWAS and *cis*-eQTL signal if posterior probability for one shared causal variant (PP4) > 0.25.

Allelic heterogeneity of gene expression in *cis* is widespread [7], and it violates the assumption of one causal variant per trait. Thus, we also used the development version of coloc [11] (https://github.com/chr1swallace/coloc/tree/condmask) in a wider 2Mb region centered at the lead ANA_C2_V2 COVID-19 GWAS variant. The enhanced version of coloc allows conditioning and masking to overcome one single causal variant assumption. For eQTL data sets from the eQTL Catalogue, we used `method = mask` with LD data from the 1000 Genomes Project [23] CEU population that matches the genetic ancestry of the population in majority of the studies in the eQTL Catalogue. For eQTL data sets from the GTEx Project, we used `method = cond` with LD calculated from the individuals that had gene expression data in the given tissue in GTEx. We used the `mode = iterative` and allowed for a maximum of three variants to condition/mask. We set the r^2^ threshold to 0.01 to call two signals independent when masking, and we set the *P*-value threshold to 10^−4^ to call the secondary signal significant. We used `method = single` for the ANA_C2_V2 COVID-19 GWAS.

As a result, the maximum PP4 (posterior probability for shared causal variants) estimates with conditioning/masking were similar to PP4 estimates from the standard coloc run (**Additional file 2: Table S1**), suggesting no additional colocalization events with secondary independent *cis*-eQTLs.

### Regional association plots for the 3p21.31 locus

Data points in the locuszoom-like regional association plots are colored by the LD between the lead ANA_C2_V2 COVID-19 GWAS variant 3:45867022:C:G and other variants in the region. For plotting data for COVID-19 GWAS and eQTLs from the eQTL Catalogue, we used genotype data from the 1000 Genomes Project, and calculated weighted average r^2^ value based on the counts of global ancestral populations in the analysis (if multiple global ancestry populations were studied). Note that for European populations, we only included non-Finnish populations in the 1000 Genomes Project. For plotting data for eQTLs from the GTEx project, we used genotype data available via dbGaP, accession phs000424.v8, and calculated r^2^ using the set of individuals that had expression data from the given tissue.

## Data Availability

The datasets analyzed during the current study are available from the following repositories: CRISPR screen data can be accessed from GEO repository under an accession number GSE158298, summary statistics of the COVID-19 GWAS (release 3) from the COVID-19 Human Genetics Initiative webpage, eQTL summary statistics can be downloaded from the eQTL Catalogue and the GTEx Portal.

https://www.ncbi.nlm.nih.gov/geo/query/acc.cgi?acc=GSE158298

https://www.covid19hg.org/results/r3/

https://www.ebi.ac.uk/eqtl/

https://gtexportal.org/

## Declarations

### Ethics approval and consent to participate

Not applicable

### Consent for publication

Not applicable

### Availability of data and materials

The datasets analyzed during the current study are available from the following repositories: CRISPR screen data can be accessed from GEO repository under an accession number GSE158298 (https://www.ncbi.nlm.nih.gov/geo/query/acc.cgi?acc=GSE158298), summary statistics of the COVID-19 GWAS (release 3) by the COVID-19 Human Genetics Initiative are available from https://www.covid19hg.org/results/r3/, eQTL summary statistics can be downloaded from the eQTL Catalogue (https://www.ebi.ac.uk/eqtl/) and the GTEx Portal (https://gtexportal.org/).

### Authors’ contributions

S.K., Z.D., N.E.S., and T.L. designed the study. Z.D., T.X.J., B.R.t., and N.E.S. designed and performed the genome-scale CRISPR screen. S.K. performed the eQTL and GWAS integration analyses. T.L. and N.E.S. supervised the work. S.K. and T.L wrote the manuscript. Z.D. and N.E.S. contributed to editing of the manuscript.

## Acknowledgements

We thank the COVID-19 Human Genetics Initiative for making the results from meta-analyses public.

