## Additional file 1 for "Integrative approach identifies *SLC6A20* and *CXCR6* as putative causal genes for the COVID-19 GWAS signal in the 3p21.31 locus"

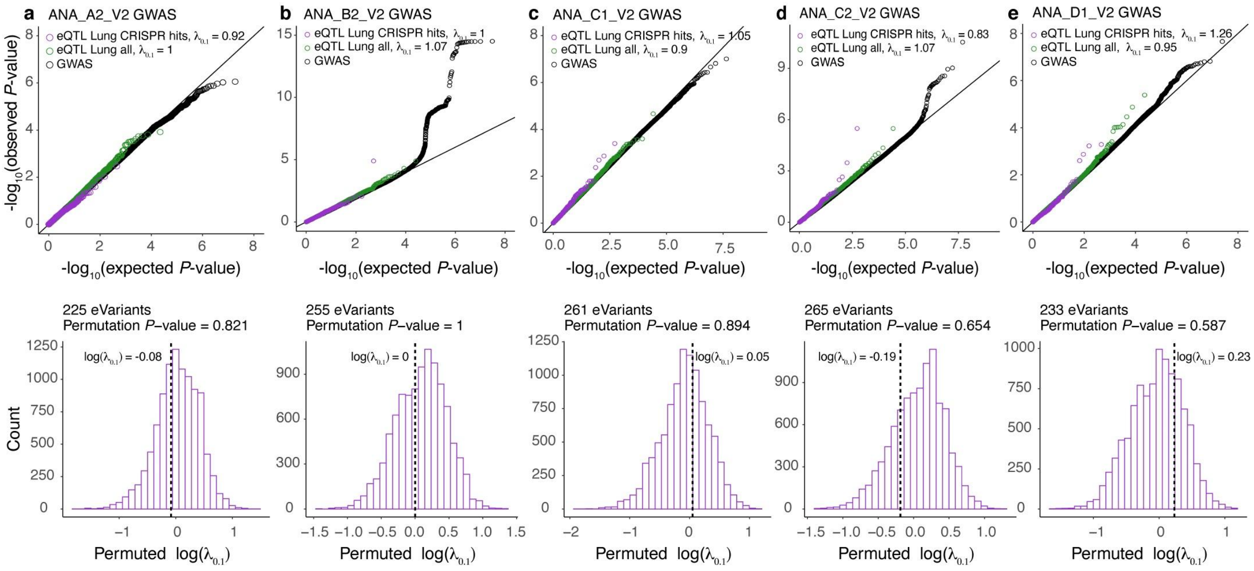


**Figure S1. Inflation of COVID-19 GWAS signal for GTEx Lung eQTLs for CRISPR screen hit genes**. The top panels shows the Q-Q plots comparing the expected and observed *P*-value distribution of COVID-19 GWAS *P*-value ($-\log_{10}$) for all the variants tested in GWAS (black), variants that are lead eQTLs in GTEx Lung (green), variants that are lead eQTLs in GTEx Lung for the top-ranked genes enriched in at least one CRISPR screen (purple) for (**a**) ANA_A2_V2 (very severe respiratory confirmed covid vs. population), (**b**) ANA_B2_V2 (hospitalized covid vs. population), (**c**) ANA_C1_V2 (covid vs. lab/self-reported negative), (**d**) ANA_C2_V2 (covid vs. population), and (**e**) ANA_D1_V2 (predicted covid from self-reported symptoms vs. predicted or self-reported non-covid) COVID-19 GWAS, respectively. Inflation estimate $\lambda_{0.1}$ measures the inflation of test statistics relative to the chi-square quantile function of 0.1. i.e., 10% of the most significant tests. The bottom panel shows the histogram of the permuted $\log(\lambda_{0.1})$ estimate to test the significance of the inflation of variants that are eQTLs for CRISPR screen hit genes in GTEx Lung in the given COVID-19 GWAS. Vertical dashed line denotes the observed $\log(\lambda_{0.1})$ value.

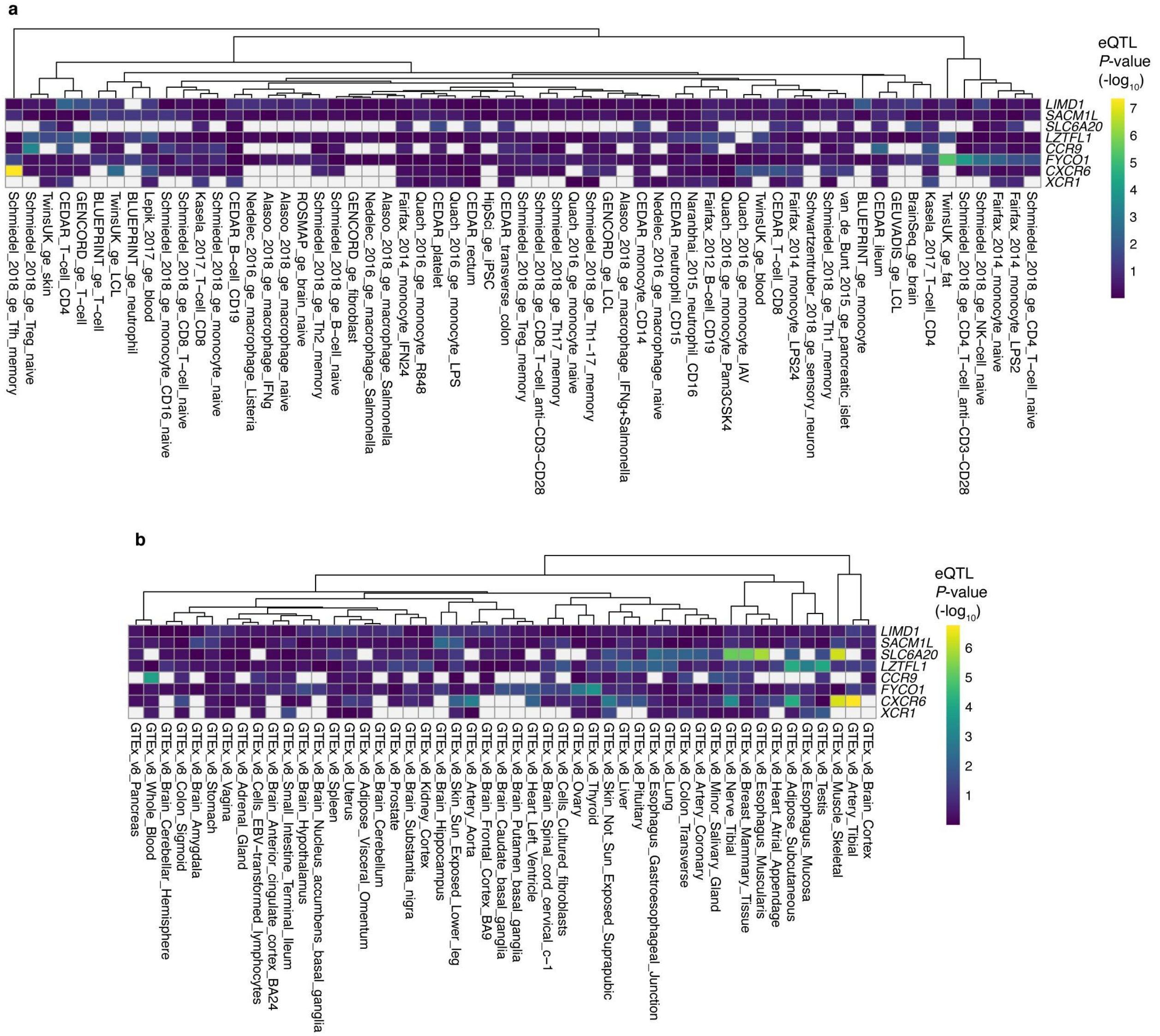


**Figure S2**. **Lead GWAS variant as an eQTL for the eight genes in the 3p21.31 locus associated with COVID-19 GWAS**. Heatmaps showing the eQTL *P*-value of the lead variant 3:45867022:C:G from COVID-19 vs population GWAS (ANA-C2-V2) for the eight genes in the 3p21.31 locus in 61 cells types and tissues from the eQTL Catalogue (**a**) and 49 tissues from GTEx v8 (**b**). The cell is colored in white, if there was no data for the variant-gene pair in the given eQTL data set (e.g., the gene was not tested in eQTL mapping). eQTL data sets are clustered using the Euclidean distance and complete linkage method.

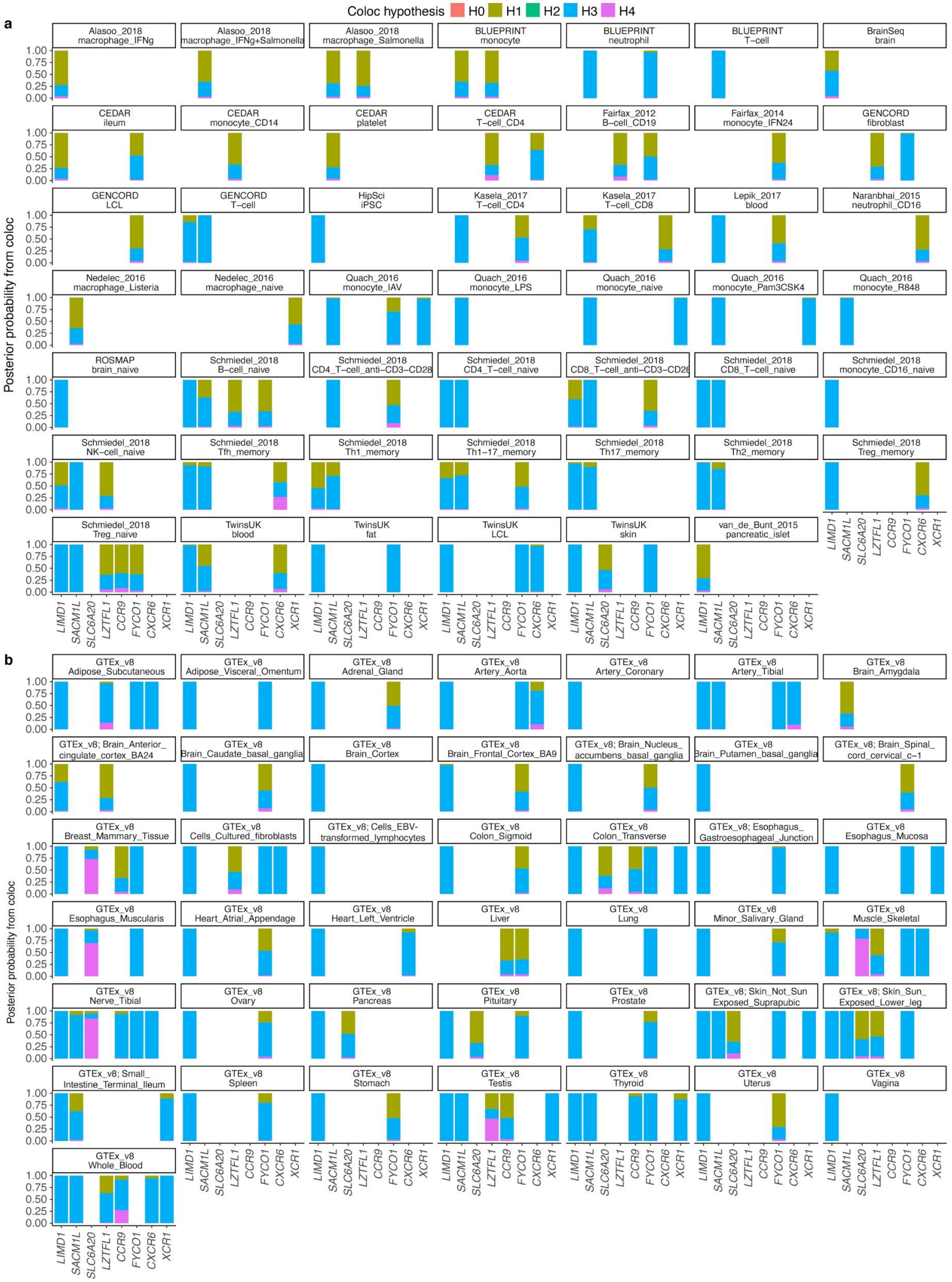


**Figure S3**. **Summary of the colocalization analysis of COVID-19 GWAS (ANA-C2-V2 - covid**

**vs. population) and eQTLs in different cell types and tissues from the eQTL Catalogue (a) and GTEx v8 (b)**. Posterior probability (*y*-axis) for the five different hypothesis H0 (no association), H1 (association in GWAS), H2 (association in eQTL), H3 (association both in GWAS and eQTL, two independent SNPs), H4 (association both in GWAS and eQTL, one shared SNP) from coloc is shown for each of the eight genes (*x*-axis) used in the colocalization analysis. For eQTL studies using microarray technology with multiple probes mapping to the same gene, we chose the probe which resulted in the lowest eQTL *P*-value for the lead COVID-19 GWAS variant to represent the gene. Note that while we observe suggestive signal for colocalization with eQTLs for *CXCR6* in Tfh memory cells (Schmiedel et al. 2018), most posterior support is attributed to H1 indicating insufficient power. Also, there is suggestive signal for colocalization with eQTLs for *LZTFL1* in Testis in GTEx, but the eQTL signal is not statistically significant (FDR > 0.05) and the lead eQTL variant has GWAS *P*-value = 0.4011.
